# Communication of Cardiovascular Disease Risk and Prevention Strategies at the Healthy Lifestyle Centres: Reality vs Expected Quality: A Cross-Sectional Mixed-Methods Study

**DOI:** 10.64898/2026.06.01.26354657

**Authors:** Sameera Upashantha Ranasinghe, Crishal Lakshitha, Sampath Tennakoon, Lakshitha Iroshan Ranasinghe

## Abstract

**INTRODUCTION:** In the multiple-risk approach of cardiovascular disease management, communication of cardiovascular disease risk and its prevention play a significant role. In Sri Lanka, this function is conducted via Healthy Lifestyle Centres.

**METHODOLOGY:** A clinical audit was conducted to describe communication quality in 79 Healthy Lifestyle Centres. A checklist developed based on Patient-Centred Communication Tools with the support of an expert panel was used. Two trained observers independently conducted the observations while the healthcare provider at the Healthy Lifestyle Centre revealed details of cardiovascular disease risk communication and health education sessions.

**RESULTS:** The majority of Healthy Lifestyle Centres involved patients in decision-making (n = 228, 92.0%), explained patient choices (n = 230, 92.8%) and responded to patients’ interest in decision-making (n = 235, 99.2%). Most patients received a summary (n = 159, 67.1%), a follow-up plan (n = 212, 89.5%) and were communicated with in jargon-free language (n = 127, 53.6%). The majority of sessions demonstrated satisfactory use of examples (95.7%, n = 22), and responsiveness to questions (73.9%, n = 17). However, most sessions were unsatisfactory regarding provision of a follow-up plan (95.7%, n = 22), encouragement of questions (95.7%, n = 22), allowing clients to talk (87.0%, n = 20), and active listening (69.6%, n = 16).

**CONCLUSIONS:** Strengthening healthcare worker training in patient-centred communication, especially shared decision-making, active listening, and encouraging patient questions, is essential to improve cardiovascular disease risk communication and patient adherence to preventive guidelines at Healthy Lifestyle Centres.

## Background and Introduction

Non-Communicable Diseases (NCDs) are responsible for a high percentage of global deaths and disabilities, causing 41 million deaths (71% of all global deaths) yearly, of which 30% were premature. Further, Low- and Middle-Income Countries (LMICs) report 77% of the total number of deaths due to NCDs (1). The 2019 Global Burden of Disease Study illustrates that Ischaemic Heart Disease (IHD) and Stroke have been ranked among the top causes of mortality in Sri Lanka (2). According to the 2021 STEPS survey conducted in Sri Lanka, 72.5% of the population did not consume five servings of fruits and vegetables daily, 30.1% did not undertake sufficient physical activity, 15.0% were current smokers, and 17.9% were current alcohol drinkers (3).

Screening for NCDs has been identified as a significant action area in the NCD strategic framework for prevention (4,5). Screening for diabetes and cardiovascular diseases (CVD) happens mainly through Healthy Lifestyle Centres (HLCs). In these centres, CVD risk is calculated using WHO ISH CVD risk prediction charts. Thereafter, referring or managing the identified patients according to the availability of resources will be carried out by these centres (6). Understanding the quality of these service provisions is essential to determine clients’ responses to the multiple-risk approach. However, these centres are mainly conducted in peripheral areas, which can have a diversity in service delivery as against the guidelines. The core of patient management at the centre is the personal communication of CVD risk and how to adhere to lifestyle changes for minimising this risk. However, communication among healthcare personnel has been found to be fraught with deficiencies worldwide (7). Such a gap can result in non-adherence to health guidance about the complex concept of CVD risk due to incomprehension by the public (7,8).

### Objective

To describe the quality of cardiovascular risk communication in HLCs in Kurunegala District of Sri Lanka.

### Methodology

A descriptive cross-sectional mixed-methods study was conducted to describe the quality of cardiovascular risk communication in HLCs. We obtained a sample of 35 randomly selected HLCs based in hospitals and 44 HLCs based in Primary Medical Care Units (PMCUs) of the Kurunegala District of Sri Lanka. Data collection was undertaken in chosen HLCs causing minimal interruption to HLCs’ functions. For risk-communication assessment, a checklist was developed based on Patient-Centred Communication Tools (PACT) (9). The PACT was originally developed to assess risk communication by pharmacists. However, due to the comprehensive and generalisable nature of the questionnaire, the tool was selected for our purpose. All communications conducted with the client, including CVD risk divulgence and health talk, were assessed using two checklists developed based on the Patient-Centred Communication Tools (PACT) questionnaire. The checklists were developed with the consensus of an expert panel consisting of a Consultant Community Physician (CCP), a Medical Officer (MO), and a Health Education Officer (HEO) who chose the most appropriate items from the original questionnaire to adapt them to assess CVD risk communication at HLCs. The data collection was conducted by two trained data collectors (for communication assessment) using independent observation during the clinic sessions at HLCs. The two assessments were compared for any discrepancies, and the consensus obtained at the end was used for the analysis.

## Results

### Quality of Cardiovascular Disease Risk Communication

The initial assessment of CVD risk was carried out by a healthcare worker, in most instances a medical officer, on a one-to-one basis. During risk communication with individual clients, rapport building was observed. In the rapport building phase, out of 237 interviews observed (3 per HLC), the following aspects involving patients in decision-making were unsatisfactory: including patients in decision-making (n = 228, 92.0%), explaining choices (n = 230, 92.8%), and responding to patients’ interest in decision-making (n = 235, 99.2%). Only 0.8% (n = 2) of communications involved agreement on short- and long-term goals. Provision of a summary (n = 159, 67.1%) and a plan for follow-up (n = 212, 89.5%) were satisfactory in most interviews. In maintaining rapport, a high proportion of conversations were found to be lacking in encouragement of questions (n = 226, 95.4%) and active listening (n = 120, 50.6%). However, responding to questions (n = 210, 88.6%) and allowing the patient to talk (n = 205, 86.5%) were prevalent in the majority of stations. Most practitioners (n = 127, 53.6%) used technical jargon-free vocabulary but did not use appropriate examples (n = 179, 75.5%). Professionalism (n = 224, 94.5%) and confidence (n = 219, 92.4%) were satisfactory in the majority. In the skill of organising the conversation, balancing patient and healthcare provider priorities was lacking in most cases (n = 218, 92.0%) (Table 1).

### Quality of Communication at the Health Education Sessions

Health education sessions were conducted for groups of clients by a Public Health Nursing Officer or Public Health Midwife in most instances. Provision of a plan for follow-up (n = 22, 95.7%) was unsatisfactory in the majority of centres, whereas provision of a summary at the end of the health talk was adequate in a high percentage (n = 17, 73.9%) of centres. In maintenance of rapport, a high proportion of conversations were found to be lacking in encouragement of questions (n = 22, 95.7%), allowing the clients to talk (n = 20, 87.0%) and active listening (n = 16, 69.6%). However, responding to questions (n = 17, 73.9%) was satisfactory in a high proportion of stations. Most of the practitioners (n = 22, 95.7%) used technical jargon-free vocabulary and used appropriate examples (n = 22, 95.7%). Professionalism (n = 22, 95.7%) and confidence (n = 16, 69.6%) were satisfactory in the majority (Table 2).

## Discussion

### Cardiovascular Disease Risk Communication

In the majority of HLCs, there was no involvement of clients in decision-making. Since HLCs focus heavily on lifestyle modification, shared decision-making would be much more effective as it has been shown to produce better health outcomes (10). However, previous research has shown that this aspect was lacking among physicians (11). This fact is reflected in the present study’s findings. Hence, the lack of follow-up visits might be due either to clients not understanding the CVD risk concept or to not recognising the necessity of follow-up. Nevertheless, follow-up advice and a summary were provided adequately in the majority of HLCs. While follow-up advice is favourable towards better patient outcomes, there is a tendency for clients to disagree with the directions given, as they were not involved in decision-making or goal setting. Such disagreement can be detrimental as CVD risk is a concept often misinterpreted by the public (12,13). Therefore, a lack of clear understanding of CVD risk communication can lead to non-adherence to healthy lifestyle modifications. Overestimation of CVD risk by the client can create unnecessary anxiety. These problems can be prevented by clear and effective CVD risk communication.

Questions from clients were not encouraged in many consultations, and low active listening was observed in the majority of HLCs in the present study. Similarly, many clinical encounters lack encouragement for clients to ask questions (14). This leads to a lack of understanding among clients, as they do not get a chance or were actively discouraged from clarifying their doubts. Further, adding to the above behaviour by healthcare providers, CVD risk communication in the UK has been described as one-way communication rather than a discussion with the client (15). It has also been found that encouraging questions from patients can lead to higher adherence to the guidelines provided by the healthcare professional (16). Nevertheless, the doctor-patient relationship in Sri Lanka is described as paternalistic (17). Therefore, CVD risk communication by medical doctors in Sri Lanka may have even more drawbacks than in the UK, due to the authoritative nature of doctors in communication with patients. Another possible reason is the lack of risk communication skills among healthcare workers, which has also been observed in other parts of the world (18,19). This issue needs to be addressed in in-service training for HLC staff. The effective use of calculated CVD risk will be very challenging in the Sri Lankan setting if the staff have not been trained in proper risk communication.

From a positive standpoint, the use of technical jargon-free language was prevalent in most HLCs. This is notable because physicians worldwide have reportedly used medical jargon, creating a barrier to communication (20). This difference may be due to socio-cultural influences in Asia, particularly where the doctor-patient bond extends beyond a purely professional relationship. Ultimately, such simple language can convey the message of CVD risk more effectively to clients.

### Quality of Communication at the Health Education Sessions

Both in Norway and the UK, HLCs conduct individual-level counselling (21,22), but in most of the HLCs in our study, the health talk remained a low priority. The primary reason might be doctors’ lack of time due to their dual roles in the HLC and Outpatient Department (OPD), especially in PMCUs. In contrast, in other countries, HLCs are conducted mainly by allied health professionals or non-clinical staff and do not involve patient screening (23,24). Whereas in Sri Lanka, both screening and counselling occur at HLCs. Differences in socio-cultural backgrounds between countries might also be a reason, as clients’ expectations differ accordingly. However, health communication remains an essential tool for behavioural change and is a significant objective of HLCs (25). The paternalistic doctor-patient relationship referred to in the above paragraph might negatively affect two-way communication between the client and the doctor. Participation in HLCs is a crucial opportunity to break down communication barriers between the doctor and the patient and to empower the public, for which health talks and individual-level counselling are essential strategies.

There were some positive aspects of the constitution at the HLCs, such as using technical jargon-free language, using examples, and providing a summary in the health talks conducted in a few of the HLCs in this study. The use of technical jargon has been found to be a significant barrier to effective outcomes in patient encounters (26). In contrast to the present study’s findings, some offshore studies have shown frequent use of medical jargon by health staff leading to confusion among patients (Russell et al., 2020). This aspect of communication being superior in HLCs in the present study may stem from grassroots-level healthcare workers’ familiarity with their clients’ literacy levels and the stronger public relationships shaped by the country’s socio-cultural influences. However, encouragement of questions and active listening were lacking in the health talks at HLCs in the current study, which are practices that should be avoided in a health talk (27). The underlying reason may be the dominant nature of healthcare workers in their relationships with clients/patients, which has also been reported in other countries (28). This can lead to comprehension problems and non-adherence in the long run.

## Conclusions

While primary healthcare workers in Healthy Lifestyle Centres demonstrate strengths in using jargon-free language, major gaps in shared decision-making, follow-up planning, and patient engagement highlight the critical need for systematic communication skills training. This will enhance cardiovascular risk communication and public health outcomes.

## Supporting information

Ethical Clearance Letter

## Ethics Approval

All observations were conducted with written informed consent from the participants. Ethical clearance for the study was obtained from the Ethics Review Committee of the University of Peradeniya, Sri Lanka.

## Data Availability Statement

The raw data used to depict these findings are available from the corresponding author on request.

## Author Contributions

**Sameera Upashantha Ranasinghe** – Conceptualisation of research question, study design, data collection, data analysis, drafting of manuscript

**Crishal Lakshitha** – Conceptualisation, study design and data collection

**Sampath Tennakoon** – Conceptualisation, monitoring and critical review

**Lakshitha Iroshan Ranasinghe** – Data analysis, drafting of work, editing, and critical review

