## Supplemental Table 1 and 2 for "Communication of Cardiovascular Disease Risk and Prevention Strategies at the Healthy Lifestyle Centres: Reality vs Expected Quality: A Cross-Sectional Mixed-Methods Study"

**Tables**

**Table 1**

***Quality of Cardiovascular Disease Risk Communication***

| **Component of communication** | **Subcomponent** | **Satisfactory**  **N = 237**  **n (%)** | **Unsatisfactory**  **N = 237)**  **n (%)** |
| --- | --- | --- | --- |
| Create rapport | Address patient appropriately ◦ Mr./Ms./Mrs. or asks how patient prefers to be addressed | 127(53.6) | 110(46.4) |
|  | Offer warm greeting | 177(74.7) | 60(25.3) |
|  | Introduce self (name and role) • | 13(5.5) | 224(94.5) |
|  | Acknowledge known information about patient | 181(76.4) | 56(23.6) |
|  | Involve patient in decision-making to the extent he/she desires | 19(8.0) | 218(92.0) |
|  | Offer and explain options and choices when appropriate ◦ | 17(7.2) | 220(92.8) |
|  | Respond to patient’s degree of interest in decision-making | 2(0.8) | 235(99.2) |
|  | Identify if there are any decision/support partners | 2(0.8) | 235(99.2) |
| Determine goals with patient | Jointly agree on short and/or long-term goals | 2(0.8) | 235(99.2) |
| Complete the visit | Provide summary (verbal and/or written) doing. | 159(67.1) | 78(32.9) |
|  | Discuss plan for follow-up (including timeline) | 212(89.5) | 25(10.5) |
| Maintain rapport | Use conversation to keep patient at ease | 12(5.1) | 225(94.9) |
|  | Encourage questions | 11(4.6) | 226(95.4) |
|  | Respond to questions | 210(88.6) | 27(11.4) |
|  | Allow patient to talk ◦ Do not interrupt or cut off patient | 205(86.5) | 32(13.5) |
|  | Demonstrate attentive listening and interest in patient | 113(47.7) | 124(52.3) |
|  | Suppress own negative reactions | 117(49.4) | 120(50.6) |
|  | Acknowledge patient’s efforts toward understanding key messages, changing behaviours, and improvement | 114(48.1) | 123(51.9) |
| Appropriate language to patient’s health literacy | Use proper vocabulary and clear, jargon-free language | 127(53.6) | 110(46.4) |
|  | Use applicable analogies and/or specific examples ◦ Verbal or pictorial | 58(24.5) | 179(75.5) |
| Confidence | Confident when stating what one knows or does not know | 219(92.4) | 18(7.6) |
|  | Display confidence during encounter | 223(94.1) | 14(5.9) |
| Professionalism | Display appropriate physical appearance, comportment, demeanor, tact, manners, etc | 224(94.5) | 13(5.5) |
| Organization | Balance patient and health care provider priorities | 19(8.0) | 218(92.0) |
|  | Maintain focus | 224(94.5) | 13(5.5) |
|  | • Flexible, yet retains control of interview | 157(66.2) | 80(33.8) |

**Table 2**

***Quality of Communication at the Health Education Sessions***

| **Component of communication** | **Subcomponent** | **Satisfactory**  **N = 23**  **n (%)** | **Unsatisfactory**  **N = 23**  **n (%)** |
| --- | --- | --- | --- |
| Complete the visit | Provide summary (verbal and/or written) doing. | 17(73.9) | 6(26.1) |
|  | Discuss plan for follow-up (including timeline) | 1(4.3) | 22(95.7) |
| Maintain rapport | Use conversation to keep patient at ease | 21(91.3) | 2(8.7) |
|  | Encourage questions | 1(4.3) | 22(95.7) |
|  | Respond to questions | 17(73.9) | 6(26.1) |
|  | Allow patient to talk ◦ Do not interrupt or cut off patient | 3(13.0) | 20(87.0) |
|  | Demonstrate attentive listening and interest in patient | 7(30.4) | 16(69.6) |
|  | Suppress own negative reactions | 7(30.4) | 16(69.6) |
|  | Acknowledge patient’s efforts toward understanding key messages, changing behaviours, and improvement | 19(82.6) | 4(17.4) |
| Appropriate language to patient’s health literacy | Use proper vocabulary and clear, jargon-free language | 22(95.7) | 1(4.3) |
|  | Use applicable analogies and/or specific examples ◦ Verbal or pictorial | 22(95.7) | 1(4.3) |
| Confidence | Confident when stating what one knows or does not know | 16(69.6) | 7(30.4) |
|  | Display confidence during encounter | 16(69.6) | 7(30.4) |
| Professionalism | Display appropriate physical appearance, comportment, demeanour, tact, manners, etc | 22(95.7) | 1(4.3) |
| Organization | Balance patient and health care provider priorities | 3(13.0) | 20(87.0) |
|  | Maintain focus | 22(95.7) | 1(4.3) |
|  | Flexible, yet retains control of interview | 22(95.7) | 1(4.3) |
