## Supplementary material for "Communication of Cardiovascular Disease Risk and Prevention Strategies at the Healthy Lifestyle Centres: Reality vs Expected Quality: A Cross-Sectional Mixed-Methods Study": Ethical Clearance Letter

**Annex X**


### ETHICS REVIEW COMMITTEE
#### Faculty of Medicine
#### University of Peradeniya


---

Dr. R.A.S.U. Ranasinghe,
198/13, Wilgoda Road,
Kurunegala.

15.08.2019

Dear Dr. Ranasinghe,

**ETHICAL CLEARANCE LETTER**

The Ethics Review Committee, Faculty of Medicine, University of Peradeniya has reviewed and discussed the protocol of Research Project No **2019/EC/29** entitled **"Comparison of CVD Risk Currently Practiced with that of Revalidated Risk Prediction Tools among 30-60 Year Olds, their Socio-cultural Response to CVD Risk Prediction and the Quality of Services Related to CVD Risk Prediction in Healthy Life Style Centers in Kurunegala RDHS Area"** submitted by you on **22.05.2019.** The committee has decided to approve the **version 2.0** of the referenced protocol on **08.08.2019**, subject to the following conditions that are mandatory:-

- It is understood that the study is being conducted in Kurunegala RDHS Area.
- Any amendment or deviation to this study protocol should not be implemented until it is reviewed and approved by the ERC, Faculty of Medicine, Peradeniya. The required amendments/deviations should be submitted to the ERC, Faculty of Medicine, Peradeniya using the **Amendment Submission Form**.
- This certificate is valid until **07.08.2020**, if and when an extension is required; a properly filled **Protocol Extension Submission Form with a progress report and a justification for extension** should be submitted to the ERC, Faculty of Medicine, Peradeniya, one month before the termination date.
- Submission of a report on Serious Adverse Events (SAE) to ERC every 3rd month during the period of study.
- Any Serious Adverse Event that occurs during the conduct of the study should be reported to the ERC Faculty of Medicine, Peradeniya immediately.
- The study should be conducted after obtaining informed consent from participant/guardian.
- Submission of progress report on ethical issues to the ERC, Peradeniya at the completion of one year period. (Please use the progress report format given in the website)
- Submission of a final report to the ERC, Peradeniya at the end of the study.
- The study has to be conducted in compliance with the approved protocol; failing to oblige may terminate the approval.

Thank you

**Dr. S.T. Kudagammana**
Secretary / Ethics Review Committee

Secretary
Ethics Review Committee
Faculty of Medicine
Peradeniya

---


Prof. A. Abeyagun[illegible]
Dean, Faculty of Medicine
University of Peradeniya
Peradeniya.
TP: 0812388840

Prof.I.B. Gawarammana
Chairperson, ERC
Department of Medicine
University of Peradeniya
0812396470

Dr. S.T. Kudagammana
Secretary, ERC
Head / Department of Paediatrics
University of Peradeniya
0812396410
